# Methodological issues in visible LED therapy dermatological research and reporting

**DOI:** 10.1101/2024.09.12.24313560

**Authors:** David Robert Grimes

## Abstract

The advent of mass-market Light Emitting Diodes (LEDs) has seen considerable interest in potential dermatological applications of LED light photobiomodulation (PBM) for a range of conditions, with a thriving market for direct-to-consumer LED treatments, including red light, blue light, and yellow light wavelengths. Evidence of efficacy for many conditions is however decidedly mixed, with starkly different outcomes reported by different authors. Due to the wide range of irradiances and wavelengths used, interpretation, comparison, and even efficacy evaluation is often impossible or prohibitive, impeding evidence synthesis. This work establishes a framework for objectively cross-comparing patient dose in terms of fluence, and a model for contrasting received dose to typical solar dose at ground level to facilitate interpretation of results and evidence synthesis. This allowes direct cross-comparison of patient skin fluence from LED PMB treatments under different regimes, and a means for evidence synthesis. This was applied to LED PMB data from 27 clinical trials to examine fluences and patient-equivalent solar exposure from LED light-sources for dermatological conditions, including acne vulgaris, wrinkle-reduction, wound-healing, psoriasis severity, and erythemal index. The results of this analysis suggest that fluences, wavelengths, and solar exposure equivalent differed by orders of magnitude in he studies analysed, with effective doses often comparable to typical daily solar exposure. Better dose quantification and plausible biological justification for various wavelengths and fluences are imperative if LED therapy studies for dermatology are to be informative and research replicability improved.

## Introduction

In recent years the home medical treatments have exploded in popularity, with the market estimated to be worth $56.45 billion by 2027, the majority comprised of therapeutic equipment^1^. Home dermatological treatments are especially popular with the wider public, sold with the allure of reducing expensive visits to dermatological experts for a range of conditions. The advent of cheap light-emitting diodes has resulted in a huge and growing array of array of LED-based skincare treatments with FDA registered devices on the market, typically in the red-light (RL), blue-light (BL), or yellow-light (YL) portion of the visible spectrum ^2^. Collectively known as low intensity photobiomodulation (PMB) devices, the many products on offer and claims around them creates some confusion - many of these projects are not rated by the FDA for efficacy, only for safety and similarity to existing products, with the FDA clarifying that *“the mechanism of actions for PBM for different clinical indications is not fully understood. Outcomes are dependent on many factors such as wavelength of light, fluence, irradiance, pulsing parameters, and beam spot size”*^3^. This note of caution from the FDA has not stopped a roaring trade in PBMs online, with LED therapies trending repeatedly on social media with tens of millions of views^4^.

Despite many influencers selling expensive home LED PBM devices including face masks and LED arrays to an eager audience, the evidence of efficacy for many of the claims made is markedly mixed. A 2023 review^5^ of 31 studies using standardized mean differences looked at RL, BL, and YL LED therapies for several skin conditions. It reported that both RL and BL were effective for acne vulgaris, skin rejuvenation (wrinkle reduction), and psoriasis treatment, and found that all RL, BL, and YL were all effective for reducing erythema index rate, whereas there was no evidence for BL having a positive impact on wound healing. This itself is curious, given that these are ostensibly very different wavelengths at opposite ends of the visible spectrum. Such results are also in stark contrast to a 2021 review^6^ specifically of RL for acne vulgaris, which found no difference in treatment outcomes between treated and control groups.

Many of the biological mechanisms postulated for the ostensible efficacy of LED therapy remain highly speculative and poorly demonstrated, in stark contrast to treatments like ultraviolet therapy^8^or even ionizing radiation^9^ where mechanisms of action are well-understood and not contested. This is compounded by significant inconsistencies in the reporting of what actual doses patients receive in LED therapy, and vast differences in wavelengths, fluences, and regimens that make direct comparison of treatments difficult. Accordingly, it becomes difficult to compare seemingly similar therapies, and even difficult to contrast them with the equivalent solar exposure required for the same fluence. To date, there has been little work done on the aspect of quantification of dose, despite the fact that this is a vital aspect to consider when ascertaining whether observed treatments effects are comparable. Even more crucially, such quantification is vital to ensure reported results are not simply spurious findings that do not benefit patients, especially when purposed mechanisms of action remain contested.

Accordingly, this work establishes a formal way of comparing patient received dose and applies this to previous studies to ascertain potential inhomogeneity in reported doses, establishing a framework to compare all treatment doses to the equivalent solar exposure time needed to achieve fluences reported at specified wavelengths in the red light (RL), blue light (BL), and yellow light (YL) portion of the spectrum. It also derives a means to compare this to typical solar irradiance at the Earth’s surface to allow clear contextualisation of findings. Importantly, this is not intended to be a systematic review of these studies, and instead uses them as a convenience sample to illustrate the inherent difficult and nuance of the problem. This work confines itself to studies directly employing LEDs for treatment, rather than any intense pulsed light or laser therapies. Equally, photodynamic therapies (PDT) involving the activation of a chemical agent are not considered in this work, though some of the dosimetric considerations discussed here may apply.

## Methods

### Analysis of included studies

Studies included in both systematic reviews^5,6^ which considered only LED therapies were assessed for information pertaining to condition treated, wavelength/s used, exposure time (*t*_*L*_) and device details. A further relevant study not included in these works due to later publication (2023-2024) was identified on Pubmed and included here. Total fluence in Joules per square centimetre per treatment, *D*, was extracted when directly reported, while for studies reporting irradiance (*E*_*L*_, Watts per square centimetre), fluence was calculated by *D* = *E*_*L*_*t*_*L*_. When all quantities were reported, units were checked for consistency. Information regarding number of patients treated and whether the study or its investigators were funded by light-treatment device manufacturers was also extracted. Papers were inspected to investigate whether they independently measured fluence or irradiance of the equipment used.

### Determination of spectral widths

RL, YL and BL LED devices are not monochromatic, and are typically specified with a central wavelength *λ*_*C*_. LEDs have a spectral width given by the Full Width at Half Maximum (FWHM), which for a typically normally distributed (or approximately normally distributed) LED profile is related to standard deviation by FWHM ≈ 2.355σ. Accordingly, for any central wavelength λc, the proportion of light at this wavelength is a function of the Gaussian spectral width, as shown in figure 1(a). For all included studies, wavelength and FWHM values were extracted when reported or available, and calculated from typical FWHM data for comparable modern LED sources^9^ when not. For an LED source centred at *λ*_*c*_ with total irradiance *E*_*T*_ and fluence *D*_*T*_, fluence also obeys a Gaussian distribution, and accordingly total fluence between any lower wavelengths (*λ*_*l*_) and higher wavelength *λ*_*u*_ in the band is akin to a Gaussian cumulative density function, given here by

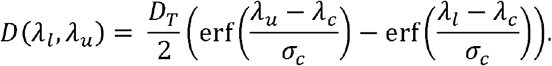

**Figure 1.**
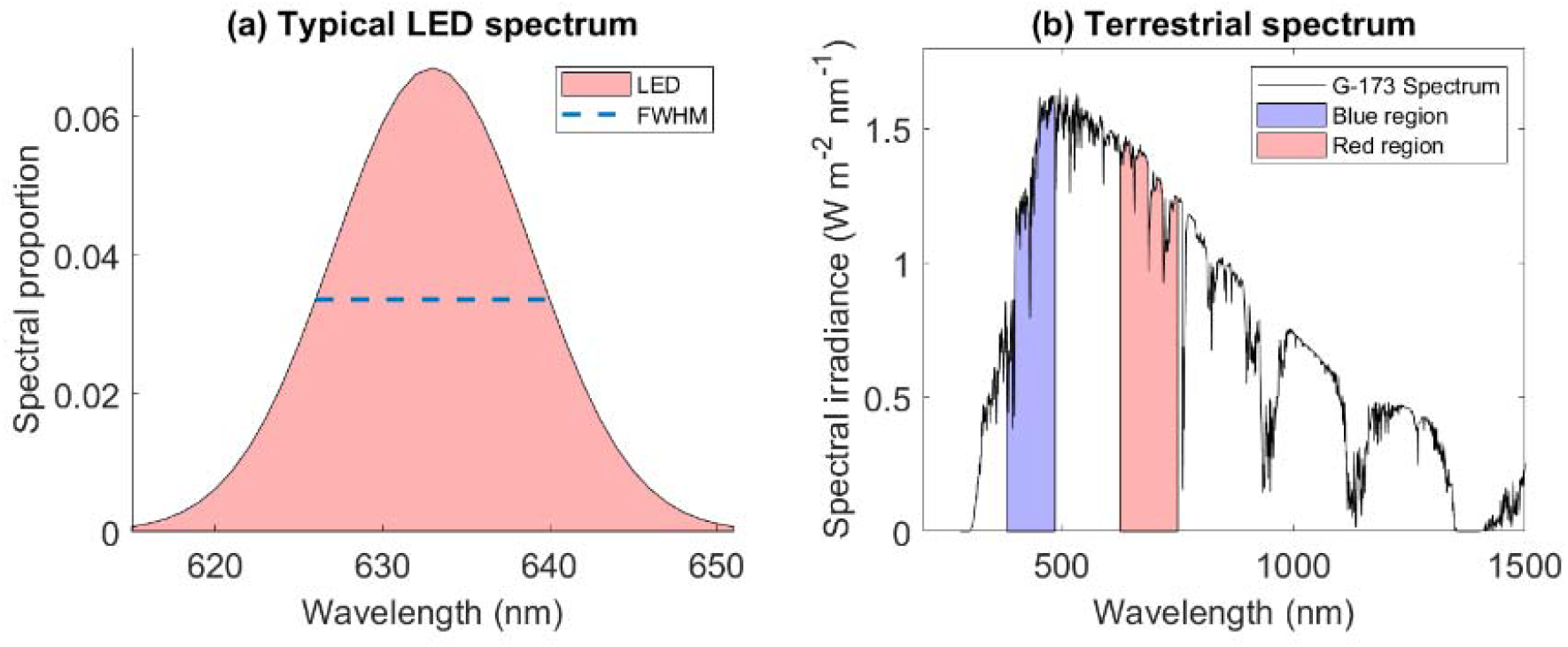
(a) Spectrum for a red LED with λ_c_ = 633 nm and FWHM = 14 nm, (b) Terrestrial solar spectrum with violet / blue band (380 - 485 nm) and red band (625 - 750 nm) highlighted

### Comparison to solar spectra exposure

The American Society for Testing and Materials (ASTM) G-173 spectra for terrestrial solar spectral irradiance mimics the conditions and tilt angle of the average latitude for the contiguous USA, with the receiving surface defined as an inclined plane at 37° tilt toward the equator, facing the sun, with atmospheric conditions as defined in the standard and an absolute air mass of 1.5 (solar zenith angle 48.19°s). Spectral irradiance, the irradiance per wavelength, defined as *S*(*λ*) has an integrated irradiance across the entire range of 1002.9 *W*/*m*^2^ a portion of which is shown in figure 1(b). To compare the ostensible therapeutic dose from a given RL/BL source to the equivalent solar exposure time, reported irraidances and doses for each study were extracted, as was central wavelength and FWHM for the LEDs used, with *D* calculated for each 1nm wavelength step along *λ* ± 3*σ*, accounting for over 97% of the total LED output. For each step, the integrated solar irradiance was also calculated with trapezoidal integration methods, given by

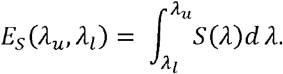

The equivalent solar exposure time, defined as the period of solar exposure required to achieve the same fluence in the same spectral band was given by

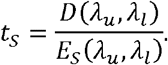

With this performed at 1nm steps between *λ*_*c*_ ± 3*σ*, the greatest value in the resultant vector (typically at central wavelength, λc) corresponded to the maximum solar exposure time required to achieve the same irradiance as a given LED treatment, facilitating direct comparison. Sample code to perform these calculations is available online at https://github.com/drg85/LEDcheck.

## Results

In the 27 LED only studies analysed, 9 (33.3%) were on acne vulgaris, 5 (18.5%) on wrinkle reduction, 3 (11.1%) on wound healing, 3 (11.1%) on psoriasis severity, and 7 (25.9%) on erythema index rate. Total number of patients ranged from 14 to 105, with a median of 26 patients. Fluences and wavelengths used in treatment differed starkly across studies, ranging from 0.1 *J cm*^−2^ − 126 *J cm*^−2^, a difference of over three-orders of magnitude. Similarly, the calculated equivalent solar time differed vastly, from 0.01-19.35 hours. Central wavelengths ranged from 405nm (BL) - 660nm (RL). None of the studies reported any dose validation procedure or independent checking, and 10 of the studies (37.0%) were sponsored by the device manufacturer, with a further 3 (11.1%) conducted by commercial dermatology practices offering the therapy under investigation as a treatment option. Histograms of the fluence and solar exposure time are shown in figure 2, and study details are given in table 1.

**Table 1.**
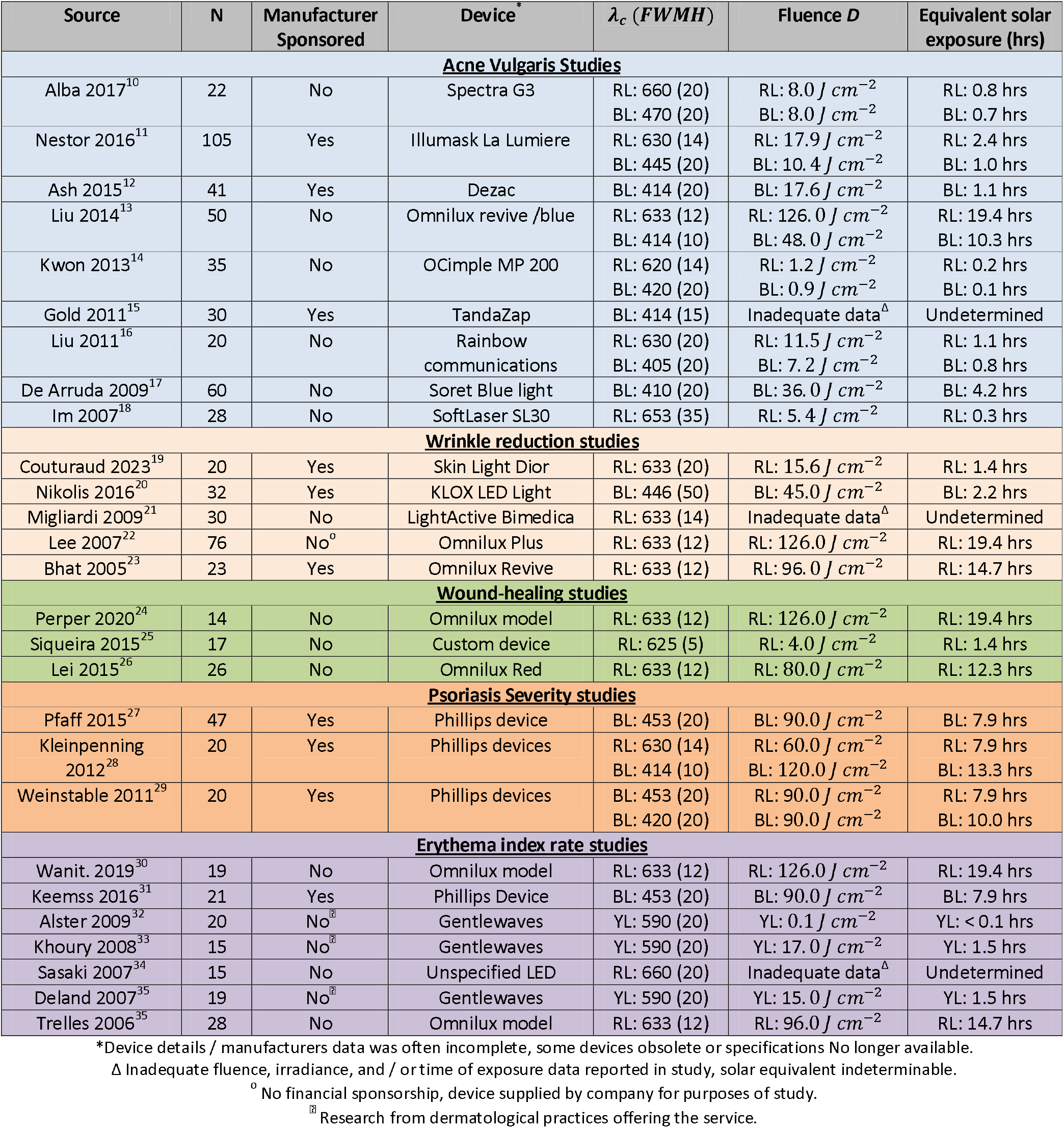
Properties of LED PBM studies analysed

**Figure 2.**
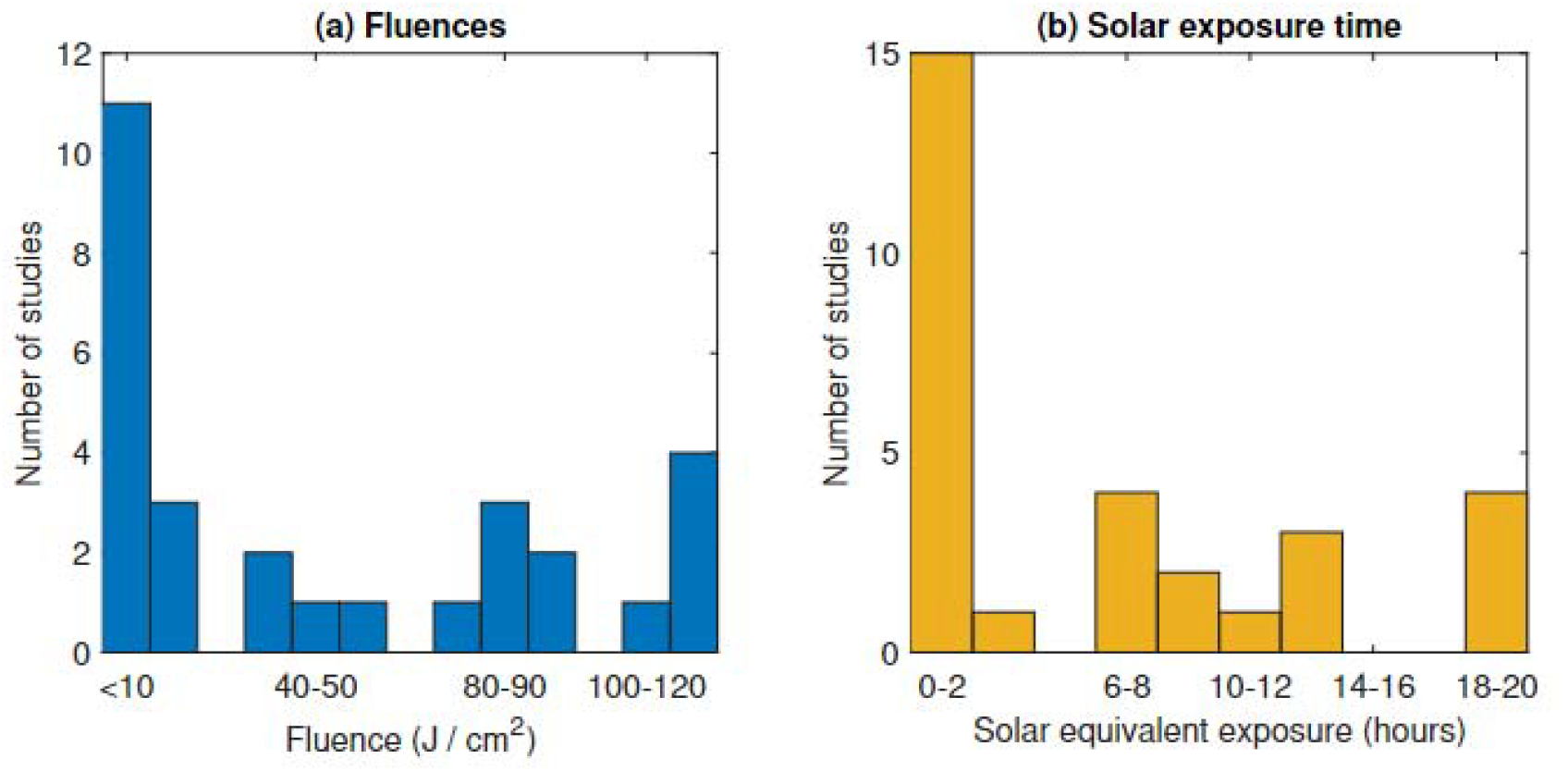
(a) Histogram of reported fluences for included studios (b) Histogram of derived equivalent solar exposure time for included studies where data was available.

## Discussion

This work outlines a useful metric for the direction comparison of dose received from LED sources in the visible spectrum to facilitate synthesis of knowledge. Equally, it shows that there is vast methodological inconsistency in experiments to date, a fundamental issue to be addressed before any deeper understanding can be garnered. In the literature analysed, there was scant justification for many of the fluences and even wavelengths used. In the former case, quoted or derived fluence rates varied by over three orders of magnitude, even for treatments ostensibly for the same condition. This was not justified in the texts, and raises serious questions over the biological rationale behind these choices. The same issue exists with respect wavelength; several of the studies reported efficacious results for both RL and BL. But these are at opposite ends of the visible spectrum, and there was scant biological discussion as to why both might be effective, nor discussion of the relatively small samples involved.

This equivalent solar exposure metric has a special utility beyond quantification of different experiments. As all the wavelengths concerned are present in the terrestrial solar spectrum, then it is important to quantify how these apparently therapeutic doses compare to normal solar exposures. In some instances, the effective exposure of a treatment was much less than the equivalent solar exposure a patient would get from under an hour of normal diffuse natural light. If LEDs therapies do have a therapeutic effect, a failure to account for this would risk confounding all exposures in both control and active arms, when even relatively minuscule amounts of normal light would deliver much more fluence at those wavelengths in some instances. It raises a crucial biological question of what actual mechanism is thought to be behind apparent benefits quoted, and whether experiments were adequately designed to answer these questions.

This work also has limitations that need to be elucidated. Critically, the solar equivalent dose calculation pivots on the fundamental assumption that the LED spectra is approximately Gaussian. This is a reasonable assumption, and most LEDs are sold with their FWHM quoted on this basis^9^. Some LEDs have minor asymmetry in their output profile at certain wavelengths^37,38^, and while this should not change estimations here, there may be situations where an LED source is for some reason non-Gaussian, in which case such calculations would be inaccurate. But equally, this raises further methodological questions about the studies considered, as few gave adequate information on source properties or reported validating the dose. It is worth noting that fluence estimates in all studies are inherently optimistic. All studies took the stated device power without clarifying whether that power referred to the device’s electrical power or optical power output, with the latter presumed in all studies. If the former was instead the case, actual fluences would be markedly lower than quoted.

It is also worth noting despite the limitations of many of these studies and the fact many are relatively old, from between 2005-2023, this has not stopped them being embraced by medical influencers and beauty bloggers online as evidence of efficacy. An abundance of influencers, some of whom have medical backgrounds, have endorsed expensive LED treatment devices claiming such studies show their efficacy, despite highly conflicting evidence. In 2024, red light therapy trended on TikTok with upwards of 70 million view^4^, and devices being offered ranging from $100 to $3500. Some of this enthusiasm is motivated by lucrative sponsorship deals, but it is most certainly exacerbated by weak studies being over interpreted without their limitations being either understood or elucidated, such as the weaknesses outlined here.

While PBM is certainly worth exploring, inadequately reported or poorly conducted studies contribute to research waste^39^, make errors harder to correct^40^, and ultimately confound the public. In biomedical science, there is dawning awareness of much literature being non-replicable, with detrimental consequences for all. Nor can meta-analysis and systematic reviews undo poor quality studies when the reporting is wildly inconsistent, as it becomes extremely difficult to compare effects. This problem is at the heart of this work, and should serve as a case study for why it is critical to compare cautiously, with many systematic reviews greatly overestimating effect size^41,42^ and themselves becoming arguably research waste. Future endeavours in this area should be cautiously reported in a consistent manner, and a biological rationale for particular fluences, wavelengths, and trials should be clearly elucidated, lest researchers end up misleading themselves and the public with spurious findings.

## Data Availability

All data available in paper.

https://github.com/drg85/LEDcheck

## Acknowledgements

DRG would like to thank Prof Enda McGlynn for his helpful discussions on LED light profiles.

## Data availability

Code to run all the analysis in this work is hosted on GitHub (https://github.com/drg85/LEDcheck) and further analytical software in various languages is available by request from the author.

## Public and Patient Involvement

As this was a cross-sectional review of published literature and methodology, It was not appropriate to involve patients in the study design or analysis at this juncture.

